# Dynamic establishment and maintenance of the human intestinal B cell population and repertoire following transplantation

**DOI:** 10.1101/2023.11.15.23298517

**Authors:** Jianing Fu, Thomas Hsiao, Elizabeth Waffarn, Wenzhao Meng, Katherine D. Long, Kristjana Frangaj, Rebecca Jones, Alaka Gorur, Areen Shtewe, Muyang Li, Constanza Bay Muntnich, Kortney Rogers, Wenyu Jiao, Monica Velasco, Rei Matsumoto, Masaru Kubota, Steven Wells, Nichole Danzl, Shilpa Ravella, Alina Iuga, Elena-Rodica Vasilescu, Adam Griesemer, Joshua Weiner, Donna L. Farber, Eline T. Luning Prak, Mercedes Martinez, Tomoaki Kato, Uri Hershberg, Megan Sykes

## Abstract

It is unknown how intestinal B cell populations and B cell receptor (BCR) repertoires are established and maintained over time in humans. Following intestinal transplantation (ITx), surveillance ileal mucosal biopsies provide a unique opportunity to map the dynamic establishment of gut lymphocyte populations. Using polychromatic flow cytometry that includes HLA allele group-specific mAbs distinguishing donor from recipient cells along with high throughput BCR sequencing, we tracked the establishment of recipient B cell populations and BCR repertoire in the allograft mucosa of ITx recipients. We confirm the early presence of naïve donor B cells in the circulation and, for the first time, document the establishment of recipient B cell populations, including B resident memory cells, in the intestinal allograft mucosa. Recipient B cell repopulation of the allograft was most rapid in infant (<1 year old)-derived allografts and, unlike T cell repopulation, did not correlate with rejection rates. While recipient memory B cell populations were increased in graft mucosa compared to circulation, naïve recipient B cells remained detectable in the graft mucosa for years. Comparisons of peripheral and intra-mucosal B cell repertoires in the absence of rejection revealed increased BCR mutation rates and clonal expansion in graft mucosa compared to circulating B cells, but these parameters did not increase markedly after the first year post-transplant. Furthermore, clonal mixing between the allograft mucosa and the circulation was significantly greater in ITx recipients, even years after transplantation, than in healthy control adults. Collectively, our data demonstrate intestinal mucosal B cell repertoire establishment from a circulating pool, a process that continues for years without evidence of establishment of a stable mucosal B cell repertoire.

## Introduction

The gastrointestinal (GI) tract is increasingly recognized as a site of abundant and diverse immune system representation and responsiveness. Additionally, immune responses local to the gut impact the entire body through systemic immune axes with the neuroendocrine system^1^. The local microbiome milieu critically shapes gut lymphocytes and immune responses^2^, with commensal and pathogenic microbes in the gut environment shaping the B cell repertoire^3,4^. The composition of gut and gut-trafficking immune cells, and their relationships with cells in other sites, continue to be catalogued and their impacts both locally and peripherally are increasingly described, including in both infectious and autoimmune disease^5,6^ as well as in transplantation^7–10^. Recently, deep immune repertoire profiling of B cell clones from adult humans established that B cell clones partition into two major networks, one in the GI tract and one in the blood, bone marrow, spleen, and lung^11^. Gut populations include many specialized B cell subsets, such as marginal zone B cells^12^ and recently reported B resident memory (BRM) cells^13^. However, how gut B cell populations and B cell receptor (BCR) repertoires are established and maintained over time in humans remains largely unknown. Such information could improve understanding of alloimmune, autoimmune and pathogen-specific immune responses in the gut.

Intestinal transplantation (ITx) provides a unique opportunity to map the dynamic establishment of gut B cell populations and BCR repertoires through the serial ileal allograft biopsies taken for clinical surveillance of rejection. Although intestinal transplants are matched for blood type in most cases, they are not matched for human leucocyte antigens (HLA), allowing discrimination of donor and recipient cells in both the periphery and within the allograft by HLA alleles^14^. ITx is used to treat patients with life threatening complications of irreversible intestinal failure resulting from congenital malformation, genetic deficiencies leading to non-function, or short bowel syndrome (e.g. massive surgical resection)^15^. Despite lymphodepleting induction therapy, high rejection rates limit transplant success and graft survival remains about 50% after 5 years^16^. Infection^17^, post-transplant lymphoproliferative disease^18^ and graft-versus-host disease (GVHD)^19,20^ also cause significant morbidity and mortality. At the time of transplant, lymphodepleting therapy reduces the recipient lymphoid mass while donor lymphocytes are transferred via the allograft at variable levels among isolated intestinal transplantation (iITx), liver-intestinal transplantation (LITx), and multivisceral transplantation (MVTx). Our previous studies suggest that the proportion of donor to recipient lymphocytes influences graft-versus-host (GvH) and host-vs-graft (HvG) reactivity, and thereby affects clinical outcomes in ITx^21–24^.

Consistently, MVTx is associated with reduced graft loss due to rejection compared to iITx^25,26^. We previously demonstrated that lymphohematopoietic graft-versus-host responses (LGVHR) frequently occur without GVHD in patients receiving MVTx in association with reduced graft rejection. Peripheral blood chimerism that includes naïve B cells is associated with engraftment of donor graft-derived hematopoietic cells in the recipient’s bone marrow and the presence of donor graft-derived GvH-reactive effector T cells that may make “space” in the marrow for the donor hematopoietic cells^22–24^. The higher donor lymphoid cell load in MVTx recipients is expected to result in stronger LGVHR than in iITx recipients. This difference may also impact B cell responses, as reduced donor-specific antibody (DSA) development is observed after liver-inclusive transplants compared with iITx^27,28^ and *de novo* DSA development is associated with increased graft loss due to rejection^29^.

Given the importance of intestinal lymphocytes in controlling infection and immune responses, intra-graft lymphocyte composition, reactivity, and function are likely important determinants of ITx outcomes. In depth analysis of intestinal T cells, hematopoietic stem and hematopoietic progenitors, and innate lymphoid cells (ILCs), including their turnover and responses in association with clinical outcomes following ITx have provided major insights into these relationships^21–24,30^. Our previous studies showed that within the allograft, while donor CD45^-^ epithelial cells were retained^22^, some ILCs persisted long term^23,30^, CD56^+^CD3^-^ NK cells turned over relatively quickly^22^, and CD3^+^ lymphocytes persisted for variable amounts of time in relation to rejection^22^ and donor age, with more rapid replacement by recipient T cells in association with rejection and younger donor age (<1 year old)^24^.

While B cells may participate in rejection of solid organ transplants both by serving as antigen-presenting cells (APCs) for T cells and by producing alloantibodies in response to T cell help^31,32^, intra-graft B cell dynamics and the maturation of B cells at both the phenotypic and clonal levels have not been analyzed in detail. We therefore performed in-depth B cell phenotyping and sequencing analysis on serial biopsies and peripheral blood samples collected after transplant, primarily in the absence of rejection or infection (during “quiescence”). Our B cell phenotyping and repertoire analyses enhance an understanding of gut mucosal B cell dynamics in relation to donor age and circulating B cell populations. These studies provide a cornerstone for future investigations on donor-reactive immune cell responses and pathologic consequences for the graft.

## Methods

### Study approval

The intestinal transplant protocol was approved by the Columbia University Institutional Review Board (IRB nos. AAAJ5056, AAAF2395, and AAAS7927). All subjects or legal guardians provided their written, informed consent and assent (Table S1). Healthy control blood and ileum tissues were obtained from deceased organ donors through a collaboration and research protocol with LiveOnNY, the organ procurement organization for the New York metropolitan area as previously described^33^ (Table S2). The use of deceased organ donor tissues is not human subjects research, as confirmed by the Columbia University IRB. Anyone outside the research group cannot reveal the identity of the study subjects by the identification (ID) numbers for patients and deceased organ donors.

### Human intestinal transplant patient recruitment and clinical protocols

Our study involves 27 intestinal transplants, including 3 re-transplants (Pt4, Pt16, Pt21) indicated as Pt4 reTx, Pt16 reTx, and Pt21 reTx. Detailed B cell phenotyping studies of 13 transplants were performed and are reported here, including Pt14, Pt17, Pt19 through Pt27, Pt4 reTx, Pt16 reTx, and Pt21reTx (Table S1). All recipients studied except Pt22 and Pt26 were pediatric (under 15 years old with a median age of 3 years at the time of transplant. Protocol and for cause endoscopic allograft biopsies were obtained as described previously^21–24^ in conjunction with clinical surveillance in the post-ITx period. Graft rejection was graded as negative, indeterminate, mild, moderate or severe based on the reported histopathologic scoring scheme^34^. Blood samples were collected up to 4 times during the first month after Tx and thereafter at least once per month when available. All patients received anti-thymocyte globulin (ATG) induction therapy (total dose: 6–10 mg/kg) followed by a maintenance regimen of long-term tacrolimus and steroids. Tacrolimus was initiated on day 1, adjusted for a target trough level of 15 to 20 ng/mL during the first month after Tx, and gradually tapered down to a maintenance level of 10 to 15 ng/mL for 2–6 months after Tx, and further tapered down to 7–10 ng/mL thereafter.

Patients received tapering doses of methylprednisolone from day 0 to day 5 after Tx, after which prednisone was maintained until indicated by the transplant team and tapered off by 6 to 12 months. Allograft rejections were treated with augmented immunosuppression based on the severity of rejection.

### Lymphocyte isolation from intestinal mucosa

We initially observed that B cells are predominately distributed in lamina propria rather than intraepithelially in the human intestinal mucosa (data not shown), consistent with observations in mice^35^. Therefore, we applied one-step Collagenase D incubation for combined intraepithelial lymphocytes (IELs) and lamina propria lymphocytes (LPLs) isolation from graft biopsy specimens, according to a protocol adapted from our previous reports^14^. Briefly, biopsy specimens were digested by stirring in flasks containing 25–50mL collagenase-containing medium (RPMI 1640, 1 mg/mL Collagenase D, 100 IU/mL penicillin-streptomycin) for 1 hour in a 37°C water bath. After passing through a 40µm filter, cells were washed with MLR media (RPMI 1640, 5% human serum, Hepes 10mM, 2-ME 50µM) before proceeding to subsequent assays. Larger full thickness surgical specimens obtained at the time of stoma closure/revision were prepared first by washing of the mucosal layer in PBS, followed by separation of the mucosal layer and isolation of IELs for other uses, prior to incubation for LPLs. For IEL isolation, minced pieces of mucosa were incubated for 30min–1hr in flasks first containing 2mmol/L dithiothreitol and 0.5 mmol/L EDTA, followed by 0.5 mmol/L EDTA, both under continuous stirring over a 37°C water bath. LPLs were isolated from the remaining tissue, digested and stirred in collagenase-containing medium as described above for 1.5–2 hours in a 37°C water bath. DNAse (0.1 mg/mL) and Fungizone (1µg/mL) were added to media when large specimens were processed. PBMCs were isolated from approximately 0.5–10mL samples of whole blood by fractionation over Histopaque gradients, followed by ACK lysis of remaining red blood cells (RBC) when necessary.

### HLA-specific cell surface staining

Candidate monoclonal HLA class I allele-specific antibodies (mAbs) were screened for the ability to discriminate donor and pretransplant (pre-Tx) recipient cells, based on clinically available molecular HLA typing information. Each HLA-specific mAb was used in combination with pan–HLA-ABC antibody and quality control tested for specificity^14^. Those that readily distinguished donor from the pre-Tx recipient PBMCs were included in lineage-specific panels of antibodies (Table S3), as reported previously^21–24^. HLA-specific antibodies used in this study are summarized as follows: HLA-ABC APC (G46-2.6, BD Biosciences, catalog 555555), HLA-ABC PE (G46-2.6, BD Biosciences, catalog 555553), HLA-ABC BV786 (G46-2.6, BD Biosciences, catalog 740982), HLA-A2/28 PE (BB7.2, BD Biosciences, catalog 558570), HLA-A2/28 FITC (One Lambda, catalog FH0037), HLA-A2/28 Biotin (One Lambda, catalog BIH0037), HLA-A9 FITC (One Lambda, catalog FH0964), HLA-A9 Biotin (One Lambda, catalog BIH0964), HLA-A3 APC (eBioscience, catalog 17-5754-42), HLA-B8 FITC (One Lambda, catalog FH0536A), HLA-B12 FITC (One Lambda, catalog FH0066), HLA-A30/31 Biotin (One Lambda, catalog BIH0067), HLA-B27 FITC (One Lambda, catalog B27F50X).

### Flow cytometry

To check B cell subset frequencies in tissue samples, cells were resuspended at 1×10^6^ cells per 100μL in FACS Buffer containing 0.1% BSA and 0.1% Sodium Azide in HBSS. Cellular staining was performed after human Fc block (Fc1.3216, BD Biosciences, catalog 564220), using combinations of the following antibodies plus HLA-specific mAbs as above, prior to DAPI staining to identify dead cells: CD3 APC-Cy7 (SK7, BD Biosciences, catalog 561800), CD14 APC-Cy7 (HCD14, BioLegend, catalog 325620), CD33 APC-Cy7 (P67.6, BioLegend, catalog 366614), CD326 APC-Cy7 (9C4, BioLegend, catalog 324245), CD19 BUV496 (SJ25C1, BD Biosciences, catalog 612938), CD20 FITC (L27, BD Biosciences, catalog 347673), CD20 APC (2H7, BD Biosciences, catalog 559776), CD38 PE-Cy7 (HIT2, BioLegend, catalog 303516), CD27-BV711 (0323, BioLegend, catalog 302833), IgM PE-CF594 (G20-127, BD Biosciences, catalog 562539), IgG V450 (G18-145, BD Biosciences, catalog 561299), IgD BV605 (IA6-2, BioLegend, catalog 348232), IgA APC (IS11-8E10, Miltenyi Biotec, catalog 130-093-073), IgA Biotin (G20-359, BD Biosciences, catalog 555884), CD21 PerCP-Cy5.5 (Bu32, BioLegend, catalog 354908), CD24 BUV395 (ML5, BD Biosciences, catalog 563818), CD45RB PE (MEM-55, BioLegend, catalog 310204), CD69 BV650 (FN50, BioLegend, catalog 310934), CD138 AF700 (MI15, BioLegend, catalog 356512), SA BUV737 (BD Biosciences, catalog 612775).

Data were acquired using an LSR II flow cytometer (BD Biosciences) with DIVA software. Analysis was carried out using FlowJo (BD Biosciences), Prism (GraphPad by Dotmatics), and Tableau (Salesforce) software.

### Cell sorting

To isolate recipient B cells from tissues, cell surface staining was performed using combinations of the following mAbs plus HLA-specific mAbs as above, after human Fc block (Fc1.3216, BD Biosciences, catalog 564220) and prior to DAPI staining to identify dead cells: CD45 V500 (HI30, BD Biosciences, catalog 560777), CD3 Per-CP-Cy5 (UCHT1, BD Biosciences, catalog 552852), CD19 BV650 (HIB19, BioLegend, catalog 302238), CD138 FITC (DL-101, BioLegend, catalog 352304), CD138 APC (DL-101, BioLegend, catalog 352308), Streptavidin PE-Cy7 (BD Biosciences, catalog 557598). Cell sorting was accomplished using an Influx cell sorter (BD Biosciences) and DIVA software. Sorted DAPI^-^ CD45^+^ CD3^-^ recipient HLA^+^ CD19^+^ and/or CD138^+^ cells were preserved in Puregene cell lysis solution (Qiagen, catalog 158906) and maintained at room temperature prior to subsequent DNA purification.

### B cell cultures and DSA Assay

For *in vitro* stimulation of B cells and plasma cells, CD19^+^ and/or CD138^+^ cells as above were sorted directly into medium containing in 10% FBS (Gemini, catalog 100-106), 2mM L-Glutamine (Gibco, catalog 25030149), 50uM 2-ME (Sigma-Aldrich, catalog M3148), 100units/mL Penicillin and 100 μg/mL Streptomycin (Gibco, catalog 15140148) in IMDM (Gibco, catalog 12440061). After washing, cells were resuspended for culture at 0.1×10^6^ cells/mL with 600 IU/mL IL-2 (R&D Systems, catalog 202IL010), 500ng/mL anti-CD40 (BioLegend, catalog 334304), 25ng/mL IL-10 (PeproTech, catalog 200-10), 2.5μg/ml CpG (Hycult, catalog HC4039), 100ng/mL IL-21 (PeproTech, catalog 200-21)^36,37^ for 7 days in an incubator controller setup maintaining 3% O_2_ (BioSpherix, catalog P360). Supernatants were concentrated approximately 20-fold using Amicon Ultra-4 Centrifugal Filters (Millipore, Catalog UFC805024). The presence of DSA in concentrated supernatants was determined by the clinical laboratory using the LabScreen Single Antigen Bead Luminex assay (One Lambda) according to the vendor’s protocol and analysis HLA Visual Software. The mean fluorescence intensity (MFI) values of beads containing donor mismatched antigens for each locus, HLA-A, HLA-B, HLA-C, HLA-DR, HLA-DP, and HLA-DQ were analyzed. For research purposes, samples in which single antigen bead IgG staining exceeded the normalized MFI baseline by 2,000 MFI (for serum) or a culture medium control by 500 MFI (for concentrated supernatants) were considered to show specificity.

### Sequencing and library preparation

Genomic DNA was extracted from FACS-sorted recipient HLA^+^ CD19^+^ and/or CD138^+^ cells from post-Tx biopsies and blood samples using the Qiagen Gentra Puregene blood kit (Qiagen, Valencia, CA, Cat. No. 158389) as previously described^11^. DNA quality and yield were evaluated by spectroscopy (Nanodrop, ThermoFisher Scientific, Waltham, MA). PCR amplification of immunoglobulin heavy-chain gene rearrangements from genomic DNA samples using primers in framework region 1 (FR1) and JH, as previously published^38,39^. DNA extraction and BCR sequencing were carried out in the Human Immunology Core Facility at the University of Pennsylvania. Experiments were performed using the Illumina 2 x 300-bp paired-end kits (Illumina MiSeq Reagent Kit v3, 600-cycle, Illumina MS-102-3003).

### Quality control and germline and clonal annotation of raw sequence data

pRESTO^40^ was used for quality control, with a protocol similar to that previously described^11,41^. Germline annotation and V and J assignment was done using IgBLAST^39^. Sequences were trimmed to IMGT position 80 (the beginning of CDR1), in order to exclude primers and imported to immuneDB. ImmuneDB^42,43^ was then used to combine sequences sharing the same VH and JH genes that had the same CDR3 length and at least 85% amino acid similarity in the CDR3. Adult sequencing data, which was used for comparisons, was taken from a previous study that used similar protocols for sequencing and quality control^11^. All data were re-annotated exactly as described above, to be comparable.

### Rejection samples

Samples that were determined to display any level of rejection, including mild or greater acute cellular rejection (ACR), or concurrence of DSA^+^ antibody-mediated rejection (AMR) via histology and serum DSA measures (MFI ≥ 2,000), were either identified specifically as in Figure 1A (associated with Figure S2) or excluded from the analyses for reasons given in the Results section.

**Figure 1.**
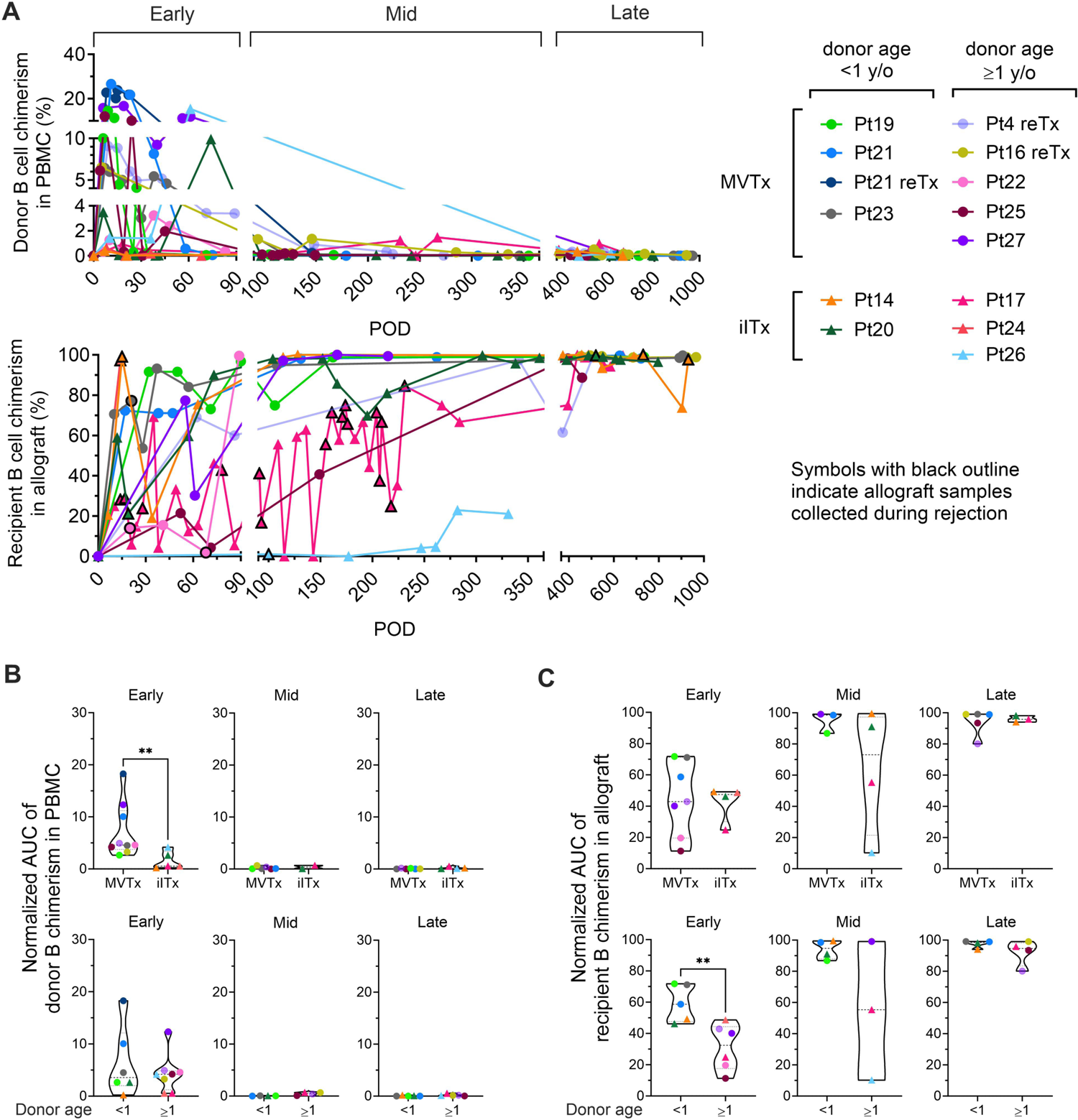
B cell chimerism in PBMCs and intestinal allografts post ITx and correlation with transplant type and donor age. (A) Chimerism of donor CD19^+^ B cells in recipient PBMC and recipient CD19^+^ B cells in ileum allograft post-Tx. Samples containing at least 40 cells in the parent gate (CD19^+^ B cells) are shown. MVTx patients are represented by circles and iITx represented by triangles. Symbols with black outline in recipient B cell chimerism in allograft plots indicate samples collected whose histology demonstrated mild to severe ACR, or concurrance of DSA^+^ AMR. Area under the curve (AUC) values normalized by days of measurement (POD_last_ – POD_first_) of donor B cell chimerism in PBMC plots (B) and recipient B cell chimerism in ileal allograft plots (C) during early (POD0–90), mid (POD91–365) and late (POD366–1000) post-Tx peroids were subgrouped by MVTx vs iITx (upper panels) and donor age <1 vs ≥1 year old (lower panels). The Mann-Whitney U test was performed to determine statistical significance (**p<0.01).

### Clone filtering

For different analyses clones were filtered by tissue type and by size. A clone was defined as detectable in a tissue if it contained at least one functional sequence (with a copy number of at least 2) in that tissue. To determine clone size within a time bracket, the number of unique instances of a clone was summed for all samples within the time bracket^38,44^. Clones were also sometimes filtered by the number of nodes observed in their lineage that were populated by a unique sequence, keeping only those clones with at least 3 nodes. Finally, we defined trunk clones as clones having 5 or more unique mutations shared by at least 85% of their unique sequences.

### Measures Reported

#### Mutation frequency

The mutation frequency of a sequence was calculated by dividing the total number of mutations found in the v gene by the sequenced length of the v gene. In our study, this metric is calculated with ImmuneDB^42,43^ and has the field name ‘v_mutation_fraction’. The average mutation frequency of a clone was calculated by averaging the v_mutation_fraction values across all unique instances in a clone. *Clumpiness:* Clumpiness^45^ quantifies how mixed two labels are within a hierarchical lineage, with a value of 0 representing labels separated from each other on different branches, and a value of 1 representing labels highly mixed together within branches. We used this metric to determine the extent of mixing between unique sequences from different tissues within lineages. To be sure our conclusions were not based on overly simple lineage structures, we only considered clones with at least 3 nodes. Clumpiness was calculated with https://github.com/DrexelSystemsImmunologyLab/clumpiness. *Diversity:* Diversity of clones was calculated with https://github.com/DrexelSystemsImmunologyLab/diversity as previously described^46^. Evenness of a repertoire was defined as the diversity of clones at order 1 normalized by their richness (the number of unique clones in a population), which, in effect, shows the diversity of clone sizes (or clonality) among a group of clones without being affected by the different number of unique clones between patients/data points.

#### Statistics

We used the nonparametric Mann-Whitney U test whenever we compared two independent samples (e.g., when comparing data points between children and adults). A log-rank (Mantel-Cox) test was performed for the Kaplan-Meier plot of freedom from moderate or severe rejection of patients with or without *de novo* Class I/II DSA in serum. When measuring changes in the same individuals between different tissues, we used the paired Wilcoxon sign-ranked test. To identify if there was a consistent direction in changes of the size of individual clones across two time points, we performed a sign test between each two time points, where each clone present at both time points that contained more unique instances at the later time bracket was given a + and each clone that contained fewer instances in the later time bracket was given a -. Clones that did not change in size were excluded from the analysis. In all cases, p-values < 0.05 were considered significant.

#### Data and materials availability

Raw BCR-seq data in FASTA format is available at Sequence Read Archive (SRA: https://www.ncbi.nlm.nih.gov/sra) under accession number PRJNA1031101. The code written in Python (version 3.5+) used to analyze BCR-seq data is available in the GitHub repository at https://github.com/DrexelSystemsImmunologyLab/Pediatric_gut_homeostatsis_paper. Requests to transfer human biospecimens outside of the organization must be submitted for Columbia’s IRB review and approval.

## Results

### B cell chimerism in tissues post ITx and correlation with type of transplant, donor age, rejection and DSA

To determine the dynamics of recipient and donor B cell populations in serial blood and allograft mucosal biopsy samples, we used surface HLA-specific staining to distinguish recipient and donor cells for each transplant (Figure S1, Table S3) in three post-Tx periods (Figure 1A): early (post operative day [POD]0–90), mid (POD91–365) and late (POD366–1000). Donor B cell chimerism was predominately detected in the blood in the early post-Tx phase, with higher peak levels and longer persistence in MVTx compared to iITx recipients, as reflected by the area under the curve (AUC) values normalized by days of measurement (POD_last_ – POD_first_) (Figure 1B). B cell chimerism declined in the mid and late post-Tx phases. These observations concur with our previous report that multilineage chimerism in blood, including B cell chimerism, is more dominant in MVTx patients^23^.

Recipient B cell replacement of donor B cells in the intestinal graft was variable and fluctuated over time. The majority of patients demonstrated consistent replacement of mucosal B cells by the recipient of at least 80% within 1 year post-Tx (Figure 1A). In the allograft, the normalized AUC of recipient B cell chimerism in the early-stage post-Tx time period was significantly greater in patients with younger (<1 year old) than older (≥1 year old) donors, regardless of the type of Tx (Figure 1C).

In contrast to observations regarding recipient T cell replacement of donor T cells in mucosal allografts^22,24^, we detected no significant correlation between the rate of recipient replacement of donor B cells in the graft and the presence of significant rejection, including mild or greater ACR and/or AMR, in the early period post-Tx (Figure S2A). The remaining analysis in our study was focused on quiescent samples because: 1) Our allograft sample collection was highly skewed towards quiescent time points in almost all patients except Pt17 (Figure 1A lower panel: symbols with black outline indicate allograft samples collected during rejection); 2) For some patients, B cell phenotyping data were only available at a few late post-Tx time points, such as Pt14 POD695 PBMC and Pt17 POD640 ileal biopsy.

We detected no significant correlation between the rate of recipient replacement of donor B cells in the graft and the development of *de novo* Class I and/or II DSA in serum in the early post-Tx period (Figure S2B). However, our data from this small cohort of patients recapitulate previous observations^27^ that *de novo* development of class I and class II DSA correlates with higher rates of moderate or severe ACR (Figure S3A). To assess the possibility that DSA might be produced within the allograft, a single mucosal sample containing sufficient B cells to permit *in vitro* culture allowed us to demonstrate local production of DSA by mucosal recipient B cells. This DSA matched the specificity detected in the serum on the same day in this patient (Pt14 POD1764), when the graft was explanted due to chronic rejection following persistent acute rejection (Figure S3B). This result suggests that recipient B cells within the allograft may give rise to DSA following ITx.

### B cell subset chimerism in the peripheral blood and allograft over time and correlation with donor age

To further characterize B cell populations after ITx, we used multicolor flow cytometry to identify naïve (CD27^-^IgD^+^) and memory (CD27^+^IgD^+/-^) subsets among recipient and donor B cells (Figures S1 and 2). From the early to the late post-Tx period, donor B cells in the PBMC were mainly naïve. Percentages of naïve cells among donor B cells were generally higher in peripheral blood than in the allograft (Figure 2A-B). Within the allograft, younger donors (<1 year old) showed significantly higher proportions of naïve B cells and significantly lower levels of memory B cells compared to older donors (≥1 year old) (Figure 2B), demonstrating B cell maturation in the human gut after infancy.

**Figure 2.**
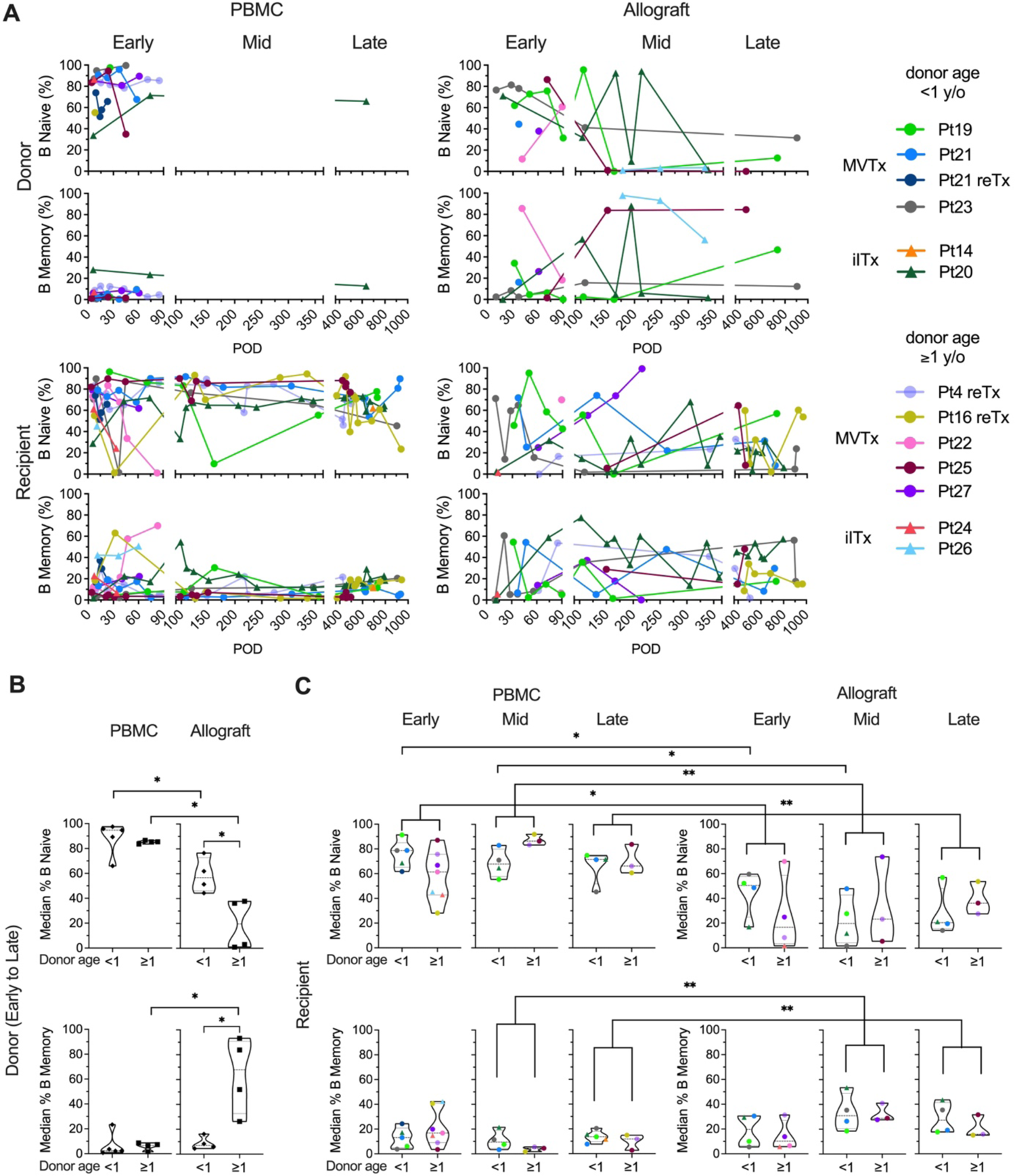
Naive and memory B cell chimerism in the PBMC and intestinal allograft over time. (A) Percentages of donor and recipient naive (CD27^-^IgD^+^) and memory B cells (CD27^+^ IgD^+/-^) in PBMC and ileal allograft during quiescence in post-Tx time brackets defined as in Figure 1. The median percentage among samples for each patient in a given period (represented by the median of the timepoints sampled) are shown for B and C. (B) Median donor naïve and memory B cell percentages among total donor B cells in PBMC and ileal allograft from early to late post-Tx periods were subgrouped by donor age. (C) Median recipient naïve and memory B cell percentages of total donor B cells in PBMC and ileal allograft during early, mid and late post-Tx peroids were subgrouped by donor age. The Mann-Whitney U test was performed to determine statistical significance (*p<0.05, **p<0.01).

Recipient B cells in the circulation and the allograft included both naïve and memory subsets, with no significant differences between younger vs older donor groups at early, middle or late post-Tx periods (Figure 2C). However, when patients with younger and older donors were combined, we found that recipient B cells within the allograft showed significantly lower median percentages of naïve cells and higher median percentages of memory cells compared to circulating recipient B cells, especially during the middle and late post-Tx periods, suggesting conversion of the initially-repopulating naïve recipient B cells or gradual entry into the mucosa of recipient memory B cells over time (Figure 2C). Even in the late stage post-Tx, median percentages of recipient B cells with the memory phenotype remained less than 50%, in contrast to donor B cells when donors were at least one year old (Figure 2B, C).

Resident memory B cells (CD69^+^CD45RB^+^) have recently been identified in human intestinal tissues^13^. Our analysis revealed the presence of donor BRM in the intestinal allograft in all post-Tx periods (Figure 3). Recipient BRM also appeared in the allograft after transplant from early timepoints in some patients and were maintained through mid to late post-Tx periods at significantly higher levels than in circulating recipient B cells (Figure 3). Recipient memory B cells in the allograft also included class-switched IgA^+^ and IgG^+^ cells at comparable levels to circulating B cells throughout the early to late post-Tx periods (Figure 3), supporting potential local secretion of DSA.

**Figure 3.**
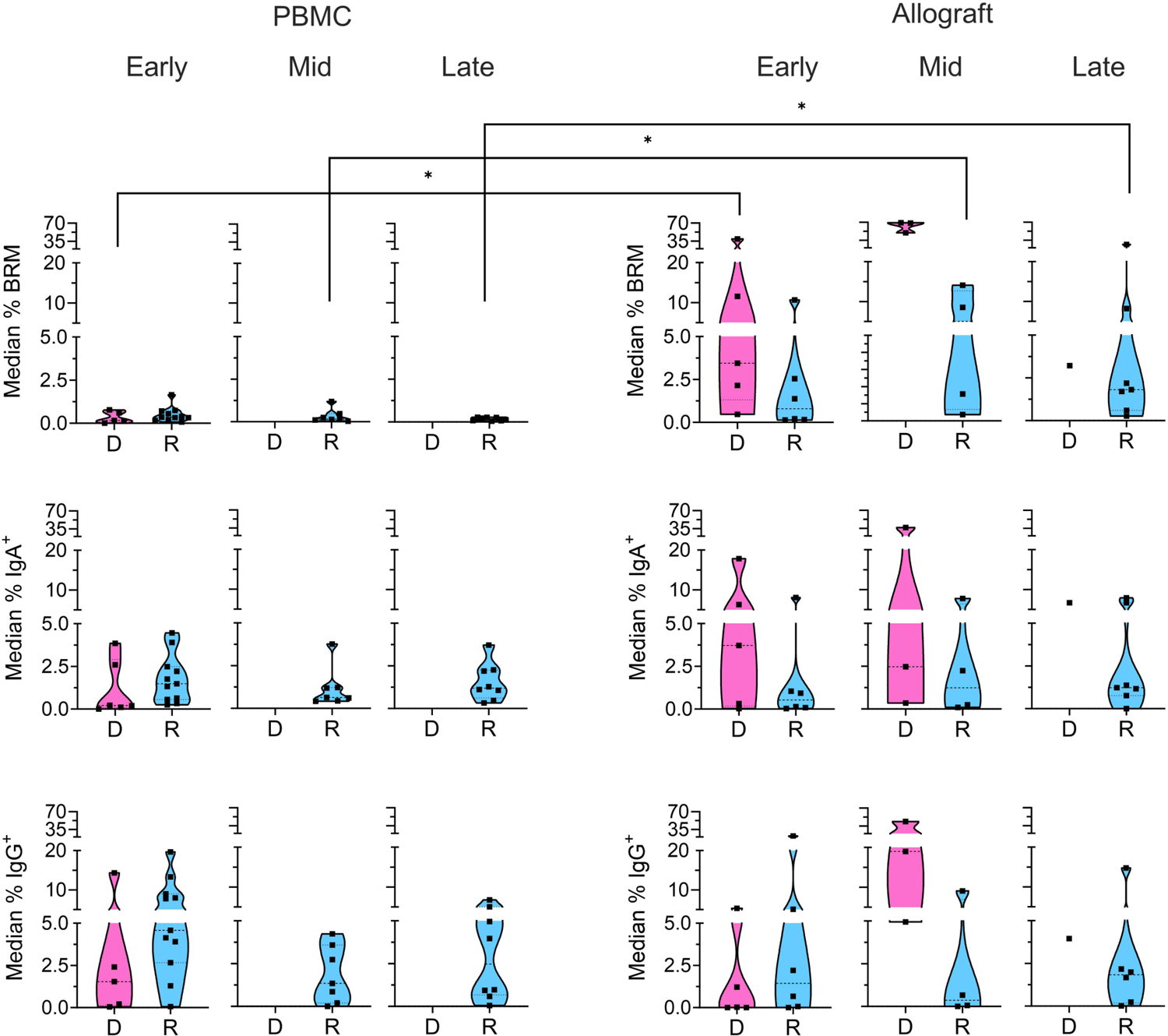
Memory subsets in recipient B cells. Median percentages of recipient BRM cells (CD69^+^CD45RB^+^), IgA^+^, and IgG^+^ B cells from the CD24^+^ memory gate (Figure S1) within total recipient B cells for each patient in the PBMC and ileal allograft during early, mid and late post-Tx peroids. In all cases, only quiescent samples exceeding a 40 cell threshold in the preceding parent gate are shown. The Mann-Whitney U test was performed to determine statistical significance (*p<0.05).

### Mutation levels of B cell clones in the blood and gut pre- and post-Tx

In order to investigate recipient B cell repertoire establishment over time in the intestinal allografts, high-throughput BCR heavy chain sequencing was performed on sorted recipient HLA^+^ B cells from serial biopsy specimens collected from pediatric ITx patients. Adult patients (Pts 22 and 26) were excluded from BCR-seq analyses due to limited post-Tx sample availability. We compared the samples from our pediatric ITx recipients to those from a cohort of deceased adult organ donors^11^. Unfortunately, the limited size biopsies obtained per timepoint did not provide sufficient numbers of BCR clones within individual post-Tx specimens to allow analysis of clonal overlap over time or between tissues for the pediatric cohort. However, we were able to evaluate the overall progression of clonal mutation (see **Methods**) by combining data from multiple time points in the early (0–90 days), middle (91–365 days) and late (>365 days) time brackets (Figure 4). Mutation rates were significantly lower in the ileal allograft specimens from pediatric recipients than in the ileum from normal adults (Figure 4A). In both cohorts, the mutation rates in circulating “blood” B cell clones were significantly lower than those in the ileum. Clones in children from pre-Tx blood samples were significantly less mutated than those detected in post-Tx blood samples (Figure 4A). For both children and adults, mutation levels in colon samples mirrored their ileum counterparts (Figure 4A).

**Figure 4.**
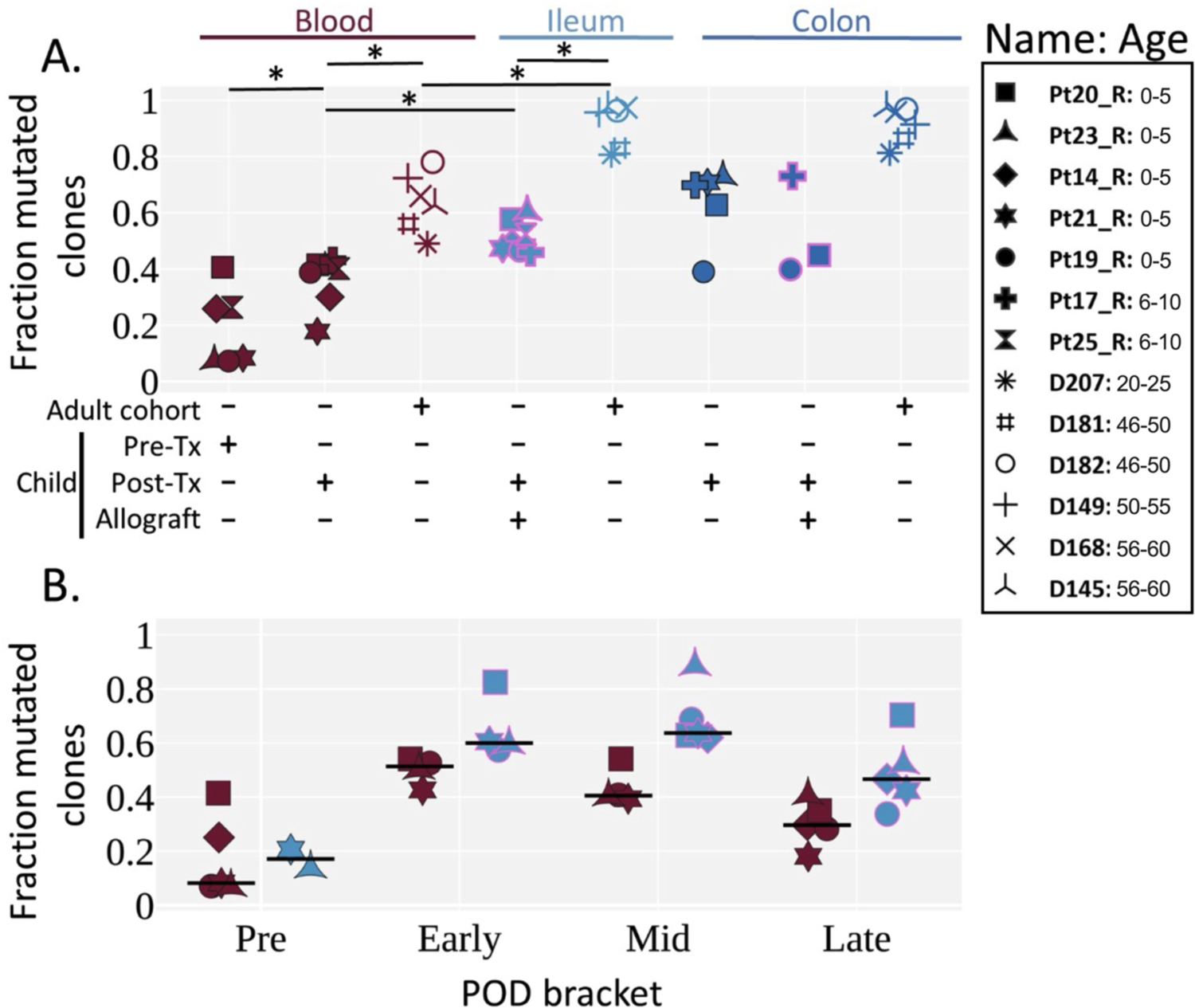
Mutation levels of BCR clones in pre- and post-Tx samples of pediatric ITx patients and adult deceased donor controls. (A) Fraction of clones with an average V gene mutation frequency >2% by tissue. Each individual is represented by a unique marker (filled markers for pediatrics, unfilled markers for adults^11^). The color of the marker represents the tissue label at the top of the graph, and the pink marker outline represents the tissue is an allograft. Pre-Tx, pretransplant; Post-Tx, post-transplant. Age ranges in years are shown. The Wilcoxon two-sided paired test was used to test for significance when the categories being compared were from the same cohort. The Mann-Whitney U test was used for significance testing between children and adults. An asterisk (*) denotes p-values <0.05. (B) Fraction of clones with an average V gene mutation frequency >2% by POD bracket for blood and ileum allograft (or native ileum for the “Pre” time bracket). Each POD is grouped into either pre, early, mid, or late time brackets (POD0, POD1–90, POD91–365 and POD>365, respectively). For each individual, the median among the PODs within a POD bracket is shown. For each POD, the samples within the POD were combined, and PODs that did not have >5 clones were omitted from the median calculation of a given POD bracket. The horizontal black line represents the median among the individuals. Tissues are colored as in Figure 4A (red for blood, light blue for ileum). (A and B) Pretransplant samples represent native recipient blood/tissue and were taken during transplantation.

Next, we looked at how clonal mutation levels changed over time (Figures 4B, S4). Differences between the ileum allograft and blood were apparent during all time brackets, although statistical significance was not reached, likely due to limited sample size. B cell clones in two available pre-Tx allograft samples (donor age <1 year old for both samples) were largely unmutated, while clones from post-Tx samples were generally more mutated, peaking in the early and middle time brackets, before demonstrating a declining trend in the late time bracket in both the allograft and the blood.

### Dynamic mapping of recipient B cell repertoire diversity in circulation and ileal allograft

To evaluate the progression of recipient B cell repertoire diversity over time, we assessed the evenness of clone distribution (see **Methods**) by time bracket (Figure 5A), which is a measure of the distribution of clonal abundances that positively correlates with clonal diversity. In some patients, the blood showed a decline in evenness in the early or middle time brackets, then returned to the high pre-Tx levels in the late time bracket. In the ileum allograft, evenness was high pre-Tx, then declined in the early and middle time brackets, before increasing in the late bracket in 4 of the 6 patients (Figure 5A). These results suggest that there were clonal expansions (i.e., increased clonality) in both the allografts and the circulating B cell pools in the first year post-Tx, but diversity eventually increased in the circulating blood clones and in the allograft in some cases during the late post-Tx period. Focusing on the 4 individuals with sequencing data whose clones were analyzed in all three time brackets, we compared how individual clones that spanned two consecutive time brackets changed in size (Figure 5B). We observed no clear trend in clone size changes between early and middle time points, whereas clones detected in the middle time bracket were significantly more likely to decrease in size in the late time bracket (Figure 5B), consistent with the increased diversity of B cell clones at this transitioning period shown in Figure 5A.

**Figure 5.**
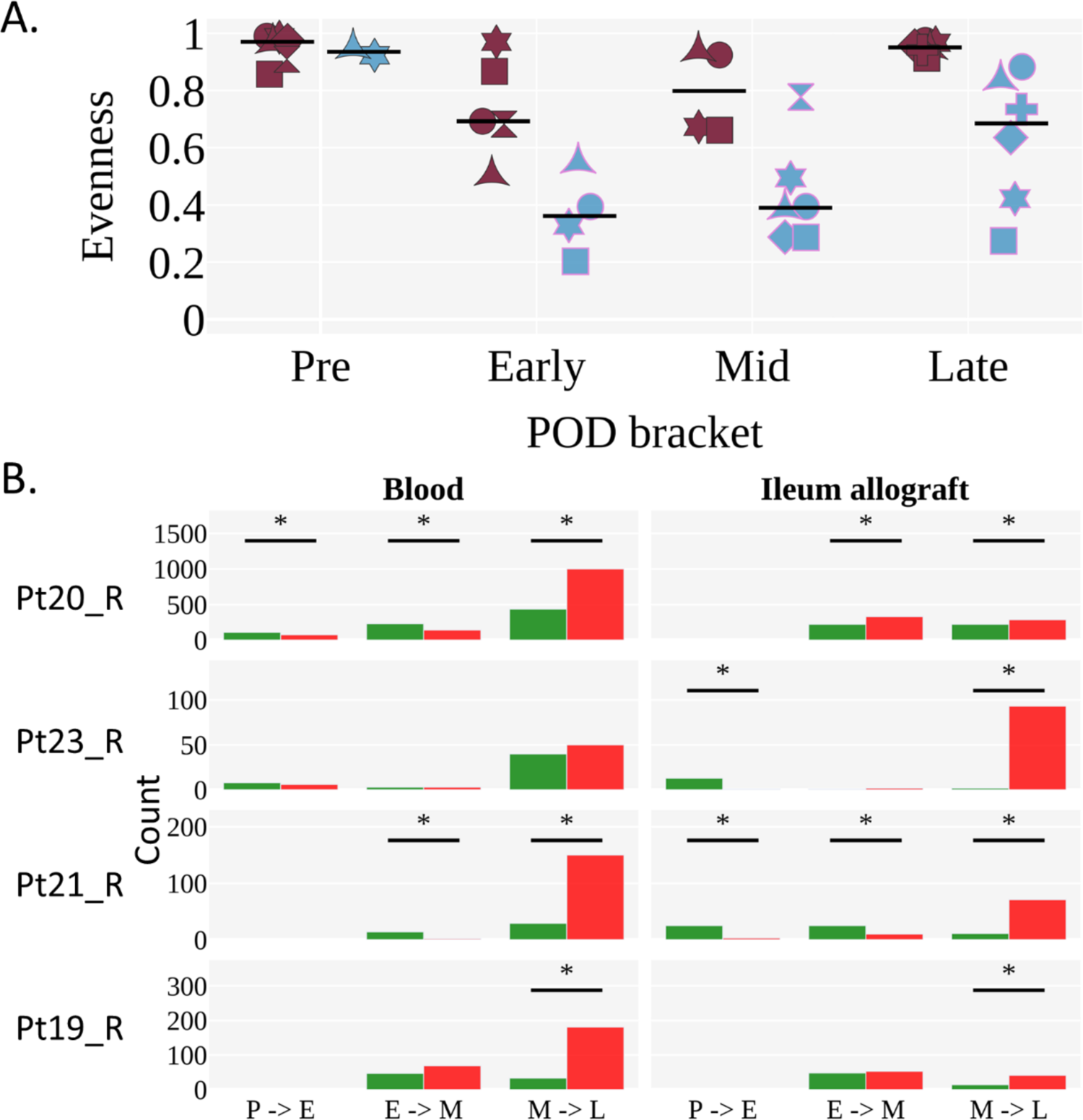
Diversity of B cell clones in peripheral blood and ileal allograft over time. (A) Evenness of clone distribution across time brackets. Evenness is calculated by dividing diversity (order 1) by richness (see **Methods**).The black horizontal line represents the median. Individuals are marked as in Figure 4. Tissues are colored as in Figure 4A (red for blood, light blue for ileum). (B) The number of clones that increase (green) or decrease (red) in size (by number of unique instances) across two time brackets. The sign test was performed to determine significant difference in the direction of clonal growth between time brackets (*p <0.05). (A and B) Each POD is grouped into either pre (P), early (E), mid (M), or late (L) time brackets, as in Figure 4B.

### B-cell clones found in pediatric blood and ileum allografts exhibit greater mixing than those in normal adults

By comparing allograft and blood specimens after combining samples from all timepoints, we were able to evaluate the extent to which clones detected in two tissues mixed or segregated their unique sequences from each tissue within their lineages. To quantify the extent to which mutations from different tissues within clonal lineages were shared along common branches and ancestors, we measured the clumpiness (see **Methods**) of mutants from different tissues in their common lineages^11,45^. Surprisingly, we observed significantly greater clumpiness between the post-Tx blood and ileum allograft in the pediatric transplant cohort compared to the low clumpiness that we had previously reported between the blood and ileum in adults^11^ (Figure 6). There was no significant difference in clumpiness between the ileum and colon of pediatric transplant recipients compared to normal adults. Given that the mutation levels of B cells in pediatric ITx patients’ gut samples tended to be lower than those in adult controls, we performed additional analyses to exclude the possibility that the higher clumpiness between blood and ileum in the pediatric transplant recipients was the result of a lack of complexity due to lack of mutations in the children’s B cell clones. We found that the clumpiness differences remained significant even when filtering for B cell clones that possess many mutations (>2% average V gene mutation) or contain a trunk of at least 5 shared mutations (Figure S5). Lastly, the mixing between the blood and ileum allograft did not appear to change with time, but rather remained at an overall comparable level throughout a majority of time points in each patient (Figure S6).

**Figure 6.**
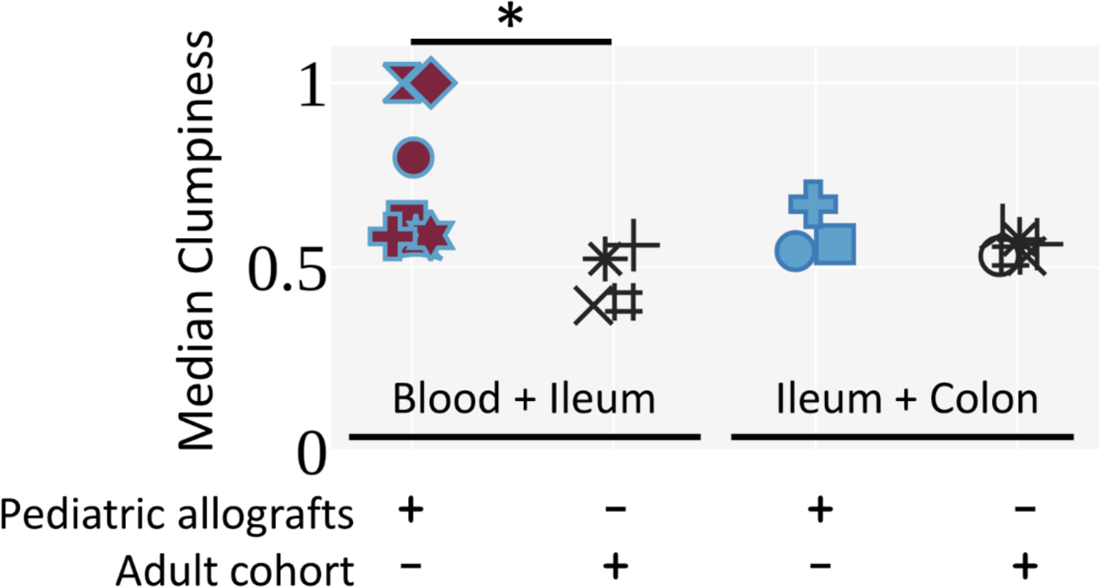
Pediatric ITx patients exhibit increased clumpiness between the blood and ileum allograft compared to adult deceased donors. Median clumpiness per individual for a given pair of tissues is shown. Clones were filtered for having 3 or more sequence nodes in their lineages (i.e., unique sequences) and being sampled in both tissues that were compared. Only medians of individuals with more than 5 clones were included. The Mann-Whitney U test was performed to determine statistical significance (*p<0.05). Individuals are marked as in Figure 4.

## Discussion

The human body is inhabited by diverse and dynamic multitudes of B cells. Although the stimuli, regulation, networks, and cellular partnerships that shape these populations are increasingly well studied, how they attain homeostasis within a tissue domain remains unknown. Here, the analysis of serial surveillance biopsies following solid organ transplantation allowed analysis of recipient B cells immigrating into the allografted intestinal mucosa, providing novel understanding of how B cell steady state is attained in human tissues. We found that recipient B cells rapidly repopulate the intestinal allograft, replacing over 80% of donor B cells within the first year post-Tx in most patients we studied. This is in contrast to T cell repopulation, where over 50% of donor CD4^+^ and CD8^+^ T cells are maintained more than 1 year in the allograft in the absence of significant rejection and with older donor age (β1 year old), but where recipient repopulation is significantly more rapid in the presence of rejection or with infant donors (<1 year old) even in the absence of rejection^22,24^. Our current study shows no correlation between the rate of recipient B cell entry into intestinal allografts and rejection. However, we did see an association of rapid recipient B cell repopulation and infant donor ileal allografts. This may be explained by the following possibilities: 1) the immature state of the infant mucosal immune system and immune cells in the gut^47–49^, requiring increased population of the mucosal compartment by circulating B cells; and/or 2) the fact that younger donors (<2 years old) have more isolated lymphoid follicles in the small intestine than older individuals^47^, resulting in increased numbers of anatomical niches for residence of recipient B cells immigrating from the circulation. The distribution pattern of B cells in the lamina propria of intestinal mucosa beyond the lymphoid follicles has not been reported for human infants. We hypothesize that this distribution may be patchy, in contrast to a more diffuse mucosal distribution pattern for T cells, on the basis of our observations from staining of CD20^+^ B cells and CD3^+^ T cells in intestinal mucosa during quiescence after ITx (data not shown, under review), and from our observations of fluctuating recipient B cell contributions to intestinal allograft biopsies taken from different sites at adjacent timepoints (Figure S1A). In contrast, recipient T cell representation in these biopsies tends to be more stable at adjacent timepoints^22,24^.

The observed detection of DSA-secreting recipient cells in the graft itself is consistent with previous reports from heart^50,51^ and kidney^52,53^ allografts, and suggests that local alloantibody production may contribute to intestinal allograft rejection, which is strongly associated with the development of DSA^27,54^. Our analysis of specificity and antibody secretion by B cells derived from the local allograft was limited by low B cell availability due to small biopsy samples and by poor survival of graft-derived B cells for study. Although we attempted multiple cell culture protocols^36,37,55^, graft-derived B cells did not survive well or secrete sufficient antibody in cultures of mucosal fragments or cell suspension cultures after sorting. This is consistent with observations that intestinal IgG-secreting cells rapidly cease antibody secretion in culture^56^.

Further investigation using a greater amount of mucosal tissue obtained from stoma revision, closure, or graft explant surgeries will be needed to strengthen this finding. Increased levels of donor B cell chimerism were detected in the peripheral blood of MVTx compared to ITx recipients, confirming previous findings^21,24^. Both donor and recipient circulating B cells were dominated by naïve phenotypes over the entire follow up period up to 1000 days post-Tx. However, in contrast to infant donor intestinal grafts, the donor B cells from older (β1 year old) intestinal grafts were predominated by memory cells. Recipient B cells entering the intestinal allograft took on the memory phenotype, including BRM phenotypes that are barely detectable in the circulation, consistent with their gut-resident memory identity.

Recipient memory B cells surprisingly maintained relatively constant frequencies (20-50%) in the mid through late post-Tx periods. In contrast to chimerism, where allografts from younger donors had faster recipient B cell replacement, there was no relationship between the naïve vs memory subset of recipient B cell frequencies in the allograft and donor age.

High throughput BCR sequencing data provided further insights into repertoire establishment in the intestinal graft over time during periods of quiescence. First, older age was associated with higher B cell mutation levels in both blood and ileum. Furthermore, B cells in the ileum showed higher mutation levels compared to those in the blood, both in pediatric ITx patients and adult deceased donor controls, similar to our earlier observations in adult organ donors^11^. Finally, we detected greater trafficking of recipient B cells between the blood and the ileum allograft in pediatric ITx patients compared to adult non-transplanted controls^11^, as demonstrated by the clumpiness measure in clonal lineages that overlapped between the blood and the ileum. While the level of trafficking remained stable over time, we tended to observe reduced evenness and diversity of the mucosal B cell repertoire in the first 3 months compared to pre-Tx blood and allograft and from 3 months to 1 year post-Tx, with some allografts reverting to pre-Tx high diversity levels in the late post-Tx period. Mutation levels in allografts rose initially post-Tx and were greater than those in the circulation, but they did not show continuous increases over time. Our findings are consistent with the possibility that a considerable fraction of B cell clones traffic continuously between the blood and the ileum allograft, rather than migrating from the blood to the ileum allograft (or vice versa) and then remaining there. While an initial surge of recipient B cell clones enters the ileum allograft in the early and middle time brackets and expand, the more uniform and evenly high diversity in the late time bracket may be explained by the possibility that many years of trafficking between the blood and ileum allograft are required post-Tx before homeostasis is achieved.

From these data we conclude that the pediatric intestinal allograft serves as an open space for recipient B cell clones to enter, and then expand and mutate. Even after 1 year, however, recipient B cell clones in the allograft revert to mutation and diversity levels comparable to what is observed early post-Tx. While we cannot rule out the possibility that recipient B cell clonal expansion within the allograft represents an immunological attack against the allograft, this seems unlikely since our repertoire analysis only included samples lacking evidence for rejection. Rather, we hypothesize that the observed recipient B-cell repertoires reflect continuous entry and turnover from the circulation into lymphoid follicle-enriched gut tissue from pediatric donors^47^. It is possible that this dynamic process echoes what occurs normally through human childhood and that steady state had not yet been established by the end of the follow-up period. Previous analyses demonstrated that the intestinal organs and lymph nodes contained a separate network of B cell clones from those in the blood, spleen and bone marrow^11^, but these analyses were performed on adult deceased donors. The differences observed here may reflect primarily the young age of our donors and recipients. If so, our data provide novel insight into the dynamic establishment of the human intestinal mucosal B cell repertoire. If, on the other hand, our results are particular to the intestinal allograft setting and do not reflect normal developmental processes, then they suggest an important failure to establish normal immune homeostasis following ITx. Studies following these patients for even longer periods, well into adulthood, will be needed to distinguish between these possibilities.

## Data Availability

All data produced in the present study are available upon reasonable request to the authors.

https://github.com/DrexelSystemsImmunologyLab/Pediatric_gut_homeostatsis_paper

https://github.com/DrexelSystemsImmunologyLab/clumpiness

https://github.com/DrexelSystemsImmunologyLab/diversity

## Acknowledgments

We thank Julissa Cabrera for assistance with the submission of the manuscript. We thank Brittany Shonts, Kryscilla Yang and Christopher Parks for their assistance with routine and after-hours sample processing and acquisition. We thank Pranay Dogra and Michelle Miron for their assistance with handoff deceased donor tissues. We thank the Flow Cytometry Core at Columbia Center for Translational Immunology (CCTI), including Siu-hong Ho and Caisheng Lu, for expertise in fluorescent panel design and cell sorting. We gratefully acknowledge the generosity of the donor families, ITx patients and their families for making this study possible.

The study is part of Program Project Grant (PPG) P01 AI106697 funded by the National Institute of Allergy and Infectious Diseases (NIAID). J.F. was supported by Nelson Faculty Development Awards from the Nelson Family Transplantation Innovation Award Program at Columbia University Irving Medical Center. E.W. was supported by NIAID training grant T32AI148099. T.H., A.S. and U.H. were also supported by the European Union’s Horizon 2020 Research and Innovation Program under grant agreement 825821 and by Israel Science Foundation (ISF) grant 1327/22. Research reported here was performed in the CCTI Flow Cytometry Core, supported in part by the Office of the Director, National Institutes of Health (NIH) awards S10RR027050 and S10OD020056 and in the Human Immunology Core at the University of Pennsylvania (RRID: SCR 022380), supported in part by NIH awards P30-AI045008 and P30-CA016520.

