## Supplemental Tables and Figures for "Dynamic establishment and maintenance of the human intestinal B cell population and repertoire following transplantation"

**Table S1. Epidemiological and clinical characteristics of ITx patients with B cell chimerism, phenotypic and sequencing data.**

SBS: Short Bowel Syndrome. NEC: Necrotizing enterocolitis. TCMR: T-cell mediated rejection. DSA: Donor-Specific Antibody

PTLD: Posttransplant lymphoproliferative disease.

| Pt (#) | Indication for Transplant | Tx Type | Recipient Age Range at Tx (years) | Recipient Sex | Donor Age Range (years) | Donor Sex | Rejection (intestinal mucosal biopsy) | GVHD | Death Or Graft Removal | *De Novo* DSA (Serum) |
|---|---|---|---|---|---|---|---|---|---|---|
| 4 reTx | Pseudo-obstruction; Berdon/MMIH syndrome | MVTx | 11-15 | Female | 0-5 | Male | No | No | No | No |
| 14 | Tufting Enteropathy | iITx | 0-5 | Male | 0-5 | Female | Early mixed rejection (mild) Late mixed rejection (Moderate) | No | Graft Removal | Early Class I, MFI >10,000, Class II MFI <6,000 Late Class I, MFI <6,000, Class II MFI >10,000 |
| 16 reTx | SBS secondary to NEC | MVTx | 0-5 | Male | 0-5 | Male | Early TCMR (mild to moderate) Late TCMR (mild) | No | No | No |
| 17 | Tufting Enteropathy | iITx | 0-5 | Male | 0-5 | Male | Early mixed rejection (mild) Mid TCMR (mild-moderate) | No | No | Early Class II MFI >6,000 |
| 19 | SBS secondary to *in utero* volvulus | MVTx | 0-5 | Male | 0-5 | Female | No | No | No | Early Class II MFI <10,000 |

| | | | | | | | | | | |
|---|---|---|---|---|---|---|---|---|---|---|
| 20 | SBS secondary to malrotation/ volvulus | iITx | 0-5 | Female | 0-5 | Female | Early mixed Rejection (mild to moderate) | No | No | Early Class I, MFI <10,000, Class II MFI <6,000 |
| 21 | Microvillus Inclusion | MVTx | 0-5 | Female | 0-5 | Male | Late TCMR (Moderate to severe) | No | Graft Removal | No |
| 21 reTx | Microvillus Inclusion | MVTx | 0-5 | Female | 0-5 | Female | Early TCMR (mild) | No | No | No |
| 22 | Budd Chiari Syndrome | MVTx | 41-45 | Female | 21-25 | Male | Early TCMR (mild) | No | Death (Sepsis) | *Preformed Class I, MFI <6,000, |
| 23 | SBS, midgut atresia | MVTx | 0-5 | Male | 0-5 | Male | Early TCMR (mild to moderate) Late TCMR (moderate to severe) | No | Death (Viral infection) | *Preformed Class I, MFI <6,000, Class II MFI <10,000 |
| 24 | SBS, cholestasis, SVC thrombosis | iITx | 6-10 | Male | 0-5 | Male | Early mixed Rejection (mild to severe) | No | Graft removal | Early Class I, MFI <10,000, Class II MFI >10,000 |
| 25 | SBS, biliary stricture, portomesenteric thrombosis | MVTx | 6-10 | Male | 0-5 | Female | Early mixed Rejection (mild) Mid mixed Rejection (mild-moderate) | No | No | Early Class I, MFI <10,000, Class II MFI >10,000 |
| 26 | HTN, SBS secondary to midgut volvulus and mesenteric ischemia | iITx | 36-40 | Female | 26-30 | Female | Early-Mid TCMR (mild to moderate) | No | No | Early Class II, MFI >2,000 |
| 27 | SBS secondary to NEC | MVTx | 0-5 | Female | 0-5 | Female | Early-Mid TCMR (mild) | No | No | No |

**Table S2. Epidemiological and clinical characteristics of adult healthy control deceased organ donors.**

| Donor (#) | Age Range (years) | Sex | Cause of Death | Tissue Usage |
|---|---|---|---|---|
| 145 | 51-55 | Male | Cerebrovascular accident | PBMC, ileum; BCR sequencing |
| 149 | 51-55 | Male | Anoxia | PBMC, ileum; BCR sequencing |
| 168 | 56-60 | Female | Cerebrovascular accident | PBMC, ileum; BCR sequencing |
| 181 | 46-50 | Male | Cerebrovascular accident | PBMC, ileum; BCR sequencing |
| 182 | 46-50 | Male | Cerebrovascular accident | PBMC, ileum; BCR sequencing |
| 207 | 21-25 | Male | Head Trauma | PBMC, ileum; BCR sequencing |
| 425 | 26-30 | Female | Anoxia | PBMC, ileum; B cell phenotyping |
| 430 | 16-20 | Male | Brain Hemorrhage | PBMC, ileum; B cell phenotyping |
| 442 | 16-20 | Male | Motor Vehicle Accident | PBMC, ileum; B cell phenotyping |
| 530 | 26-30 | Male | Cerebrovascular/stroke | PBMC, ileum; B cell phenotyping |
| 531 | 46-50 | Male | Cerebrovascular/stroke | PBMC, ileum; B cell phenotyping |

**Table S3. HLA class I typing and anti-HLA allele antibodies used to distinguish donor from recipient cells in ITx recipients.** HLA-A09 is a broad antigen HLA-A serotype that recognized the HLA-A23 and HLA-A24 serotypes. HLA-A28 is a broad antigen HLA-A serotype that recognized the HLA-A68 and HLA-A69 serotypes. HLA-B12 is a broad antigen HLA-B serotype that recognized the HLA-B44 and HLA-B45 serotypes.

| Pt (#) | Recipient HLA I type | Donor HLA type | HLA I allele-specific mABs Used to discriminate recipient from donor cells |
|---|---|---|---|
| 4 reTx | **A02** / A30<br>B42 / B53 | A68 / A74<br>B72 / B42 | Anti-HLA A02 (BB7.2) FITC<br>Anti-HLA A03 APC (First donor) |
| 14 | **A23** / A-<br>B50 / B- | **A03** / A68<br>B35 / B58 | Anti-HLA A09 Biotin<br>Anti-HLA A03 APC |
| 16 reTx | A02 / A34<br>B15 / **B44** | **A24** / 29<br>B35 / **B44** | Anti-HLA B12 FITC (First donor $B12^{-}A9^{-}$)<br>Anti-HLA A09 Biotin |
| 17 | A30 / A31<br>B40 / B53 | A24 / A32<br>**B27** / B35 | Anti-HLA B27 FITC |
| 19 | A02 / A68<br>B39 / B48 | A02 / **A03**<br>B53 / B72 | Anti-HLA A03 APC |
| 20 | A02 / **A03**<br>B50 / B52 | A02 / A68<br>**B44** / B53 | Anti-HLA A03 APC<br>Anti-HLA B12 FITC |
| 21 | A01 / A11<br>B39 / B58 | **A03** / **A03**<br>B65 / B35 | Anti-HLA A03 APC |
| 21 reTx | A01 / A11<br>B39 / B58 | **A02** / A31<br>B18 / B60 | Anti-HLA A02/A28 FITC<br>Anti-HLA A03 APC (First donor) |
| 22 | A01 / A30<br>B15 / B53 | A29 / A34<br>B27 / **B45** | Anti-HLA B12 FITC |
| 23 | **A02** / A30<br>B15 / B57 | **A03** / A24<br>B35 / B44 | Anti-HLA A03 APC<br>Anti-HLA A02/28 Biotin |
| 24 | A01 / A11 | **A68** / A74 | Anti-HLA B08 FITC |

| | | | |
|---|---|---|---|
| | **B08** / B41 | B07 /B57 | Anti-HLA A02/28 Biotin |
| 25 | **A02** / A03<br>B71 / B49 | A01 / A24<br>B08 / **B44** | Anti-HLA A02/28 Biotin<br>Anti-HLA B12 FITC |
| 26 | A01 / A33<br>B65 / B38 | A25 / A26<br>**B08** / B38 | Anti-HLA B08 FITC |
| 27 | A32 / A68<br>B35 / B57 | **A02** / **A03**<br>B07 / B51 | Anti-HLA A02 (BB7.2) FITC<br>Anti-HLA A03 APC |

**Supplemental Figure 1** Gating strategy for phenotyping B cell subsets in Pt20 POD73 PBMC (A) and POD74 ileal allograft biopsy (B) samples. HLA-specific markers distinguishing transplant donor (blue) and recipient (red) are used to identify transplant-originating donor cells in PBMC and allograft mucosa. Gates and % shown are only for recipient cells (red). Memory B cells: $CD27^{+}IgD^{+/-}$. Naïve B cells: $CD27^{-}IgD^{+}$. Transitional B cells: $CD24^{+}CD38^{+}$ among naïve B cell gating. Mature Naïve B cells: $CD24^{-/dim}CD38^{-/dim}$ among naïve B cell gating. ASC (antibody secreting cells): $CD24^{-}CD38^{+}$ among memory B cell gating. $CD24^{+}$ Memory: $CD24^{+}CD38^{-/dim}$ among memory B cell gating. BRM: $CD69^{+}CD45RB^{+}$ B cells.

**A** Pt20 POD73 PBMC

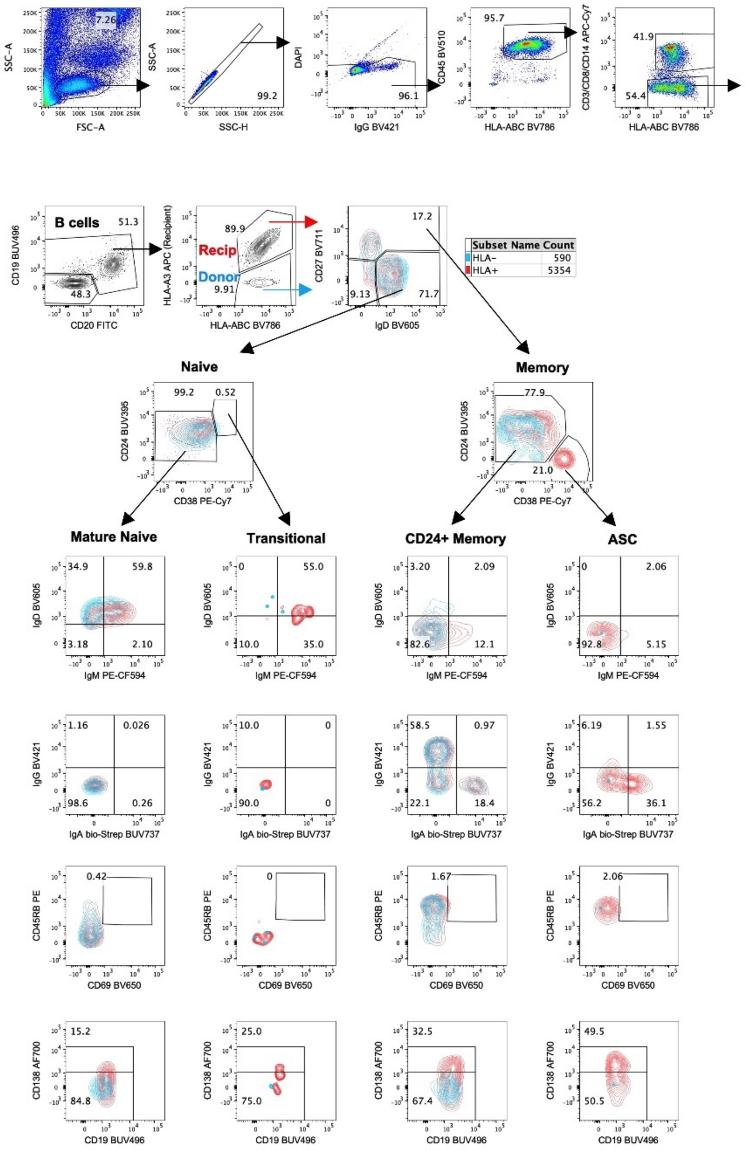


**B** Pt20 POD74 ileal allograft biopsy

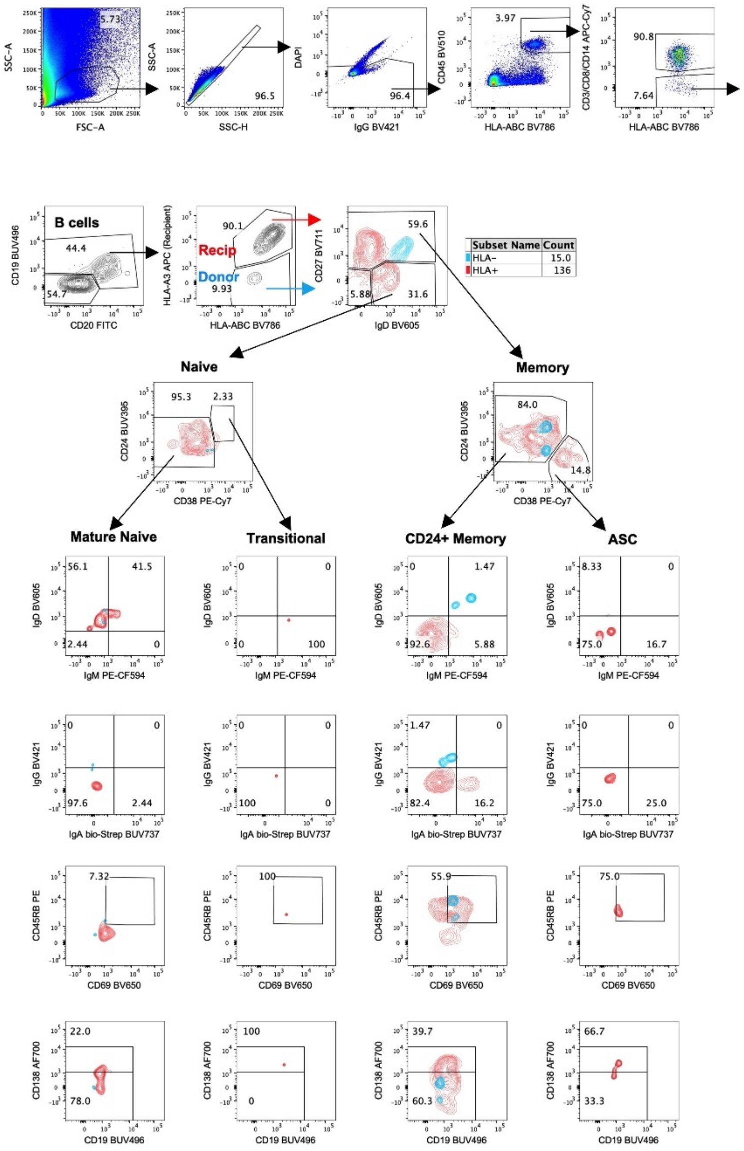

**Supplemental Figure 2** Normalized area under the curve (AUC) values of recipient B cell chimerism in allograft in patients (A) with or without acute cellular rejection (ACR) and (B) with or without *de novo* Class I/II DSA in serum during early post-Tx period (up to POD90). No significant difference was detected by Mann-Whitney U test (p>0.05).

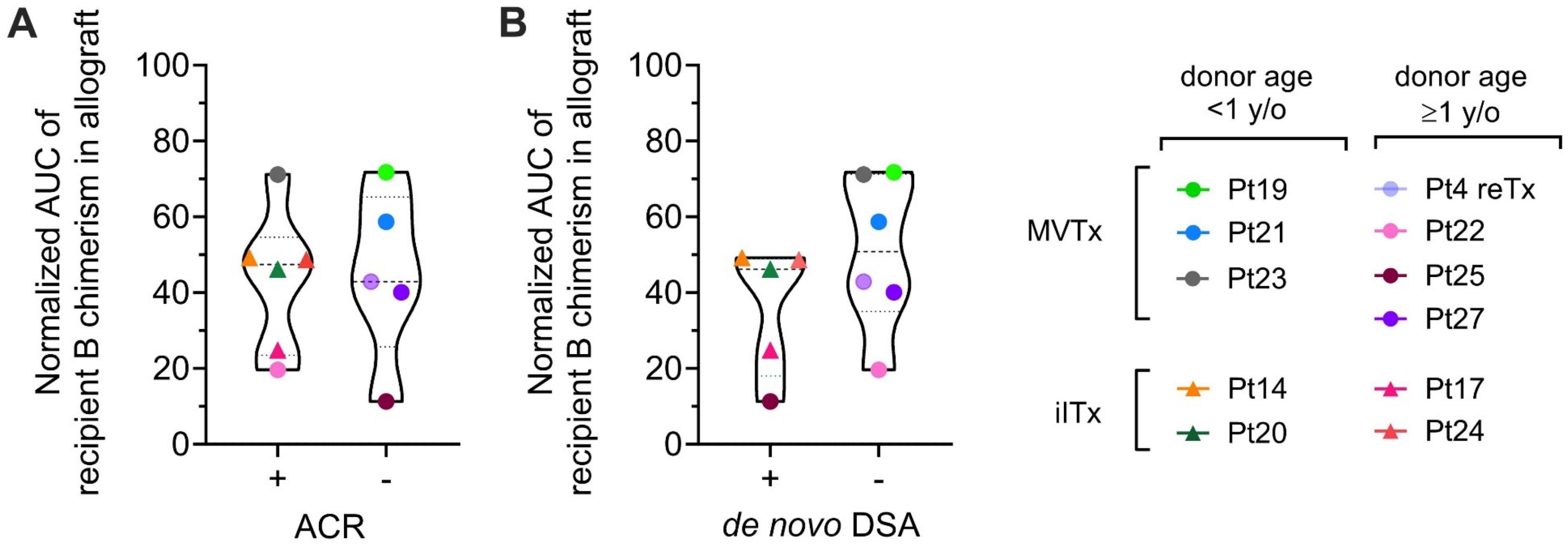

**Supplemental Figure 3** (A) *De novo* development of Class I (upper panel) and Class II (lower panel) DSA detected in post-Tx serum correlates with higher rates of moderate or severe ACR. (B) Local production of DSA by mucosal recipient B cells matched the DSA specificity detected in the serum on the same day in a patient during graft explant (Pt14 POD1764) due to chronic rejection with previously persistent ACR.

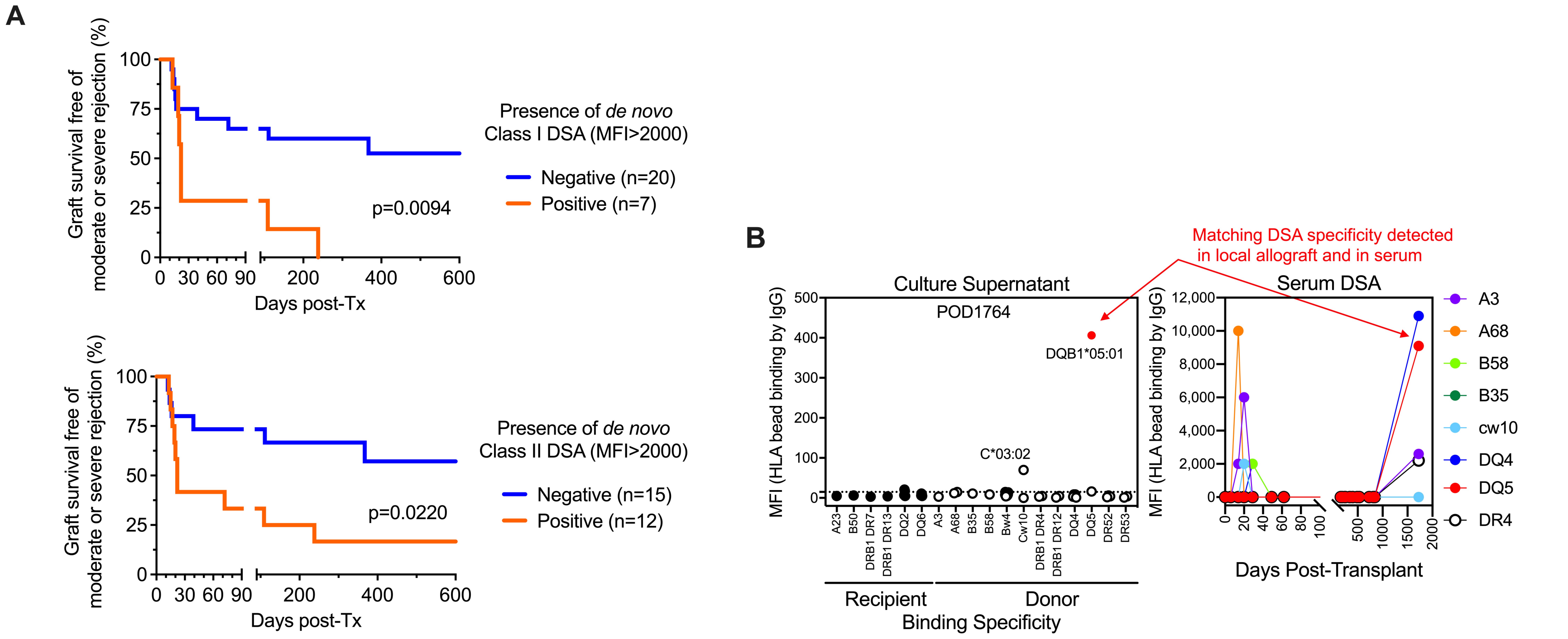

**Supplemental Figure 4.** The fraction of clones per sample with an average v gene mutation frequency >2% by POD is shown in peripheral blood and ileum allograft. The median fraction of mutated clones among adult deceased donors is shown by the dashed lines. Individuals are marked as in Figure 4. Green markers indicate pre-Tx samples (samples taken at POD0).

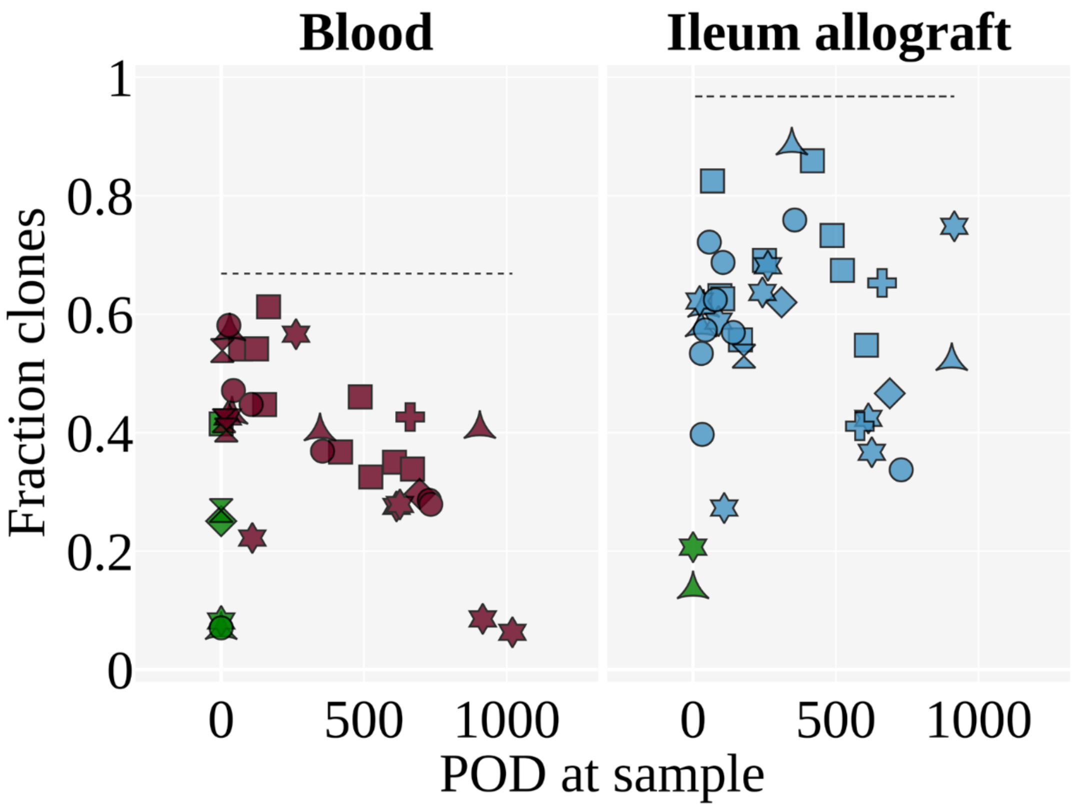

**Supplemental Figure 5.** Pediatric transplant patients exhibit increased trafficking between the blood and ileum allograft among mutated (A) and trunk (B) clones compared to adult deceased donor controls. The median clumpiness per individual for a given pair of tissues is shown. In panel (A), clones were filtered for having 3 or more sequence nodes in their lineages, being mutated (average gene mutation frequency >2%), and being sampled in both tissues that were compared. In panel (B), clones were filtered for having 3 or more unique sequences, having a trunk, and being sampled in both tissues that were compared. Only medians with greater than 5 clones were included. The Mann-Whitney U test was performed to determine statistical significance (*p<0.05). Individuals are marked as in Figure 4.

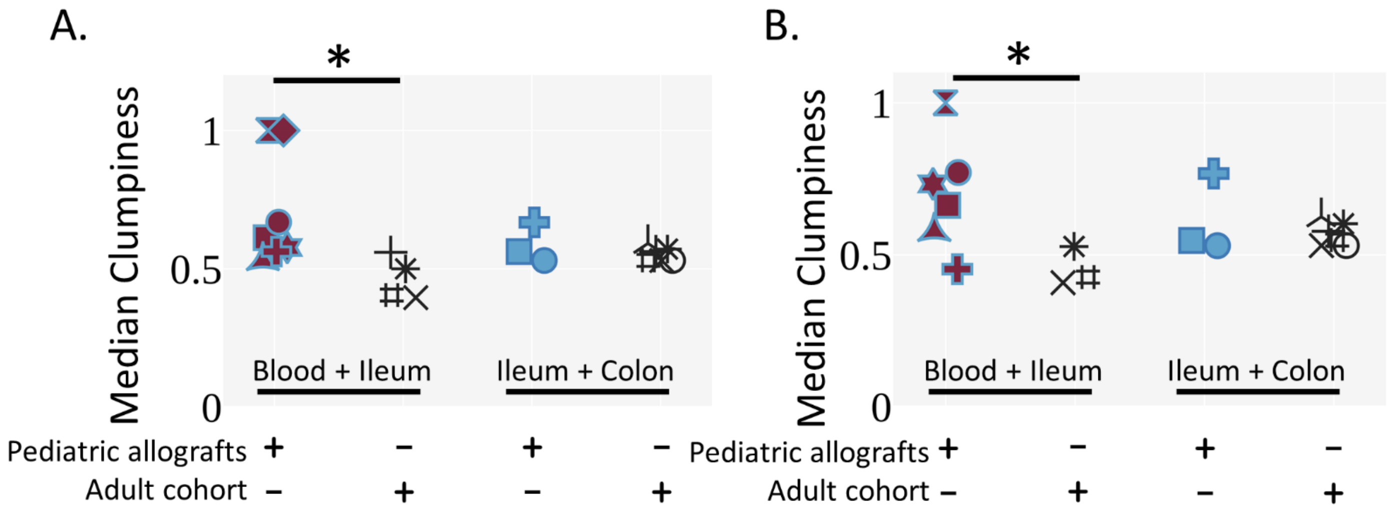

**Supplemental Figure 6.** Median clumpiness between the blood and ileum allograft (or pre-Tx ileum for POD0) per individual by POD was shown. Clones were filtered for having 3 or more unique sequences, and being sampled in both tissues that were compared. Only data points with greater than 5 clones were included. Median clumpiness between the blood and ileum tissues in adult deceased donors is shown by the dotted dashed line. Individuals are marked as in Figure 4.

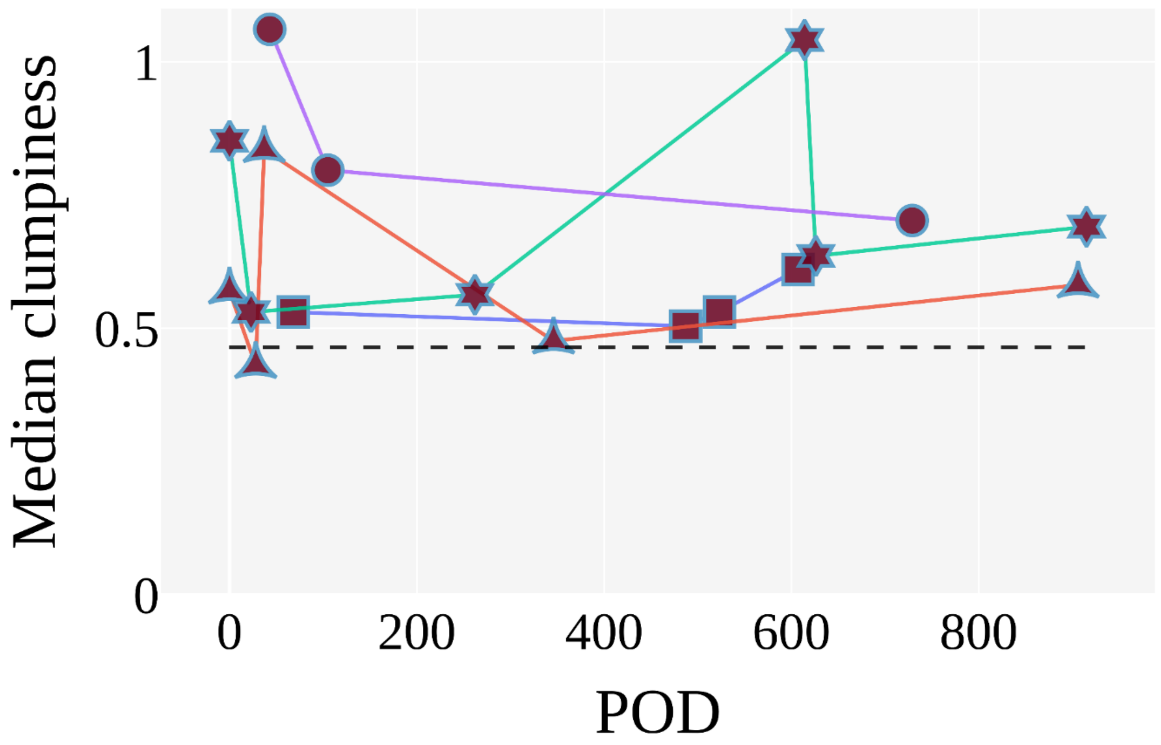
